# Disruption of spatiotemporal clustering in dengue cases by *w*Mel *Wolbachia* in Yogyakarta, Indonesia

**DOI:** 10.1101/2021.08.24.21261920

**Authors:** Suzanne M. Dufault, Stephanie K. Tanamas, Citra Indriani, Adi Utarini, Riris Andono Ahmad, Nicholas P. Jewell, Cameron P. Simmons, Katherine L. Anders

## Abstract

Dengue exhibits focal clustering in households and neighborhoods, driven by local mosquito population dynamics, human population immunity, and fine scale human and mosquito movement. We tested the hypothesis that spatiotemporal clustering of homotypic dengue cases is disrupted by introduction of the arbovirus-blocking bacterium *Wolbachia* (*w*Mel-strain) into the *Aedes aegypti* mosquito population. We analysed 318 serotyped and geolocated dengue cases (and 5,921 test-negative controls) from a randomized controlled trial in Yogyakarta, Indonesia of *w*Mel deployments. We find evidence of spatial clustering up to 300m among the 265 dengue cases (3,083 controls) in the untreated trial arm. Participant pairs enrolled within 30 days and 50m had a 4.7-fold increase (compared to 95% CI on permutation-based null distribution: 0.1, 1.2) in the odds of being homotypic (i.e. potentially transmission-related) as compared to pairs occurring at any distance. In contrast, we find no evidence of spatiotemporal clustering among the 53 dengue cases (2,838 controls) resident in the *w*Mel-treated arm. Introgression of *w*Mel *Wolbachia* into *Aedes aegypti* mosquito populations interrupts focal dengue virus transmission leading to reduced case incidence; the true intervention effect may be greater than the 77% efficacy measured in the primary analysis of the Yogyakarta trial.

## 1 Introduction

Dengue places seasonal pressure on healthcare systems and public health resources throughout the tropical and subtropical world, with an estimated 100 million cases globally each year. The disease burden is growing in both case load and countries affected [1]. The four serotypes of dengue virus (DENV) are transmitted between humans primarily by the *Aedes aegypti* mosquito, a species that thrives in urban settings where breeding sites and human blood sources co-exist in close proximity. These bionomic factors make households a primary location for DENV transmission risk and the focal nature of DENV transmission has been well observed [2, 3, 4, 5, 6].

The temporal and spatial scale at which dengue case clustering occurs is informative about both the underlying transmission dynamics and the opportunity to intervene and interrupt transmission. A prospective index-cluster study in rural Thailand [2] demonstrated highly focal DENV transmission among children residing within 100 meters and 15 days of an index case. A retrospective analysis of serotyped dengue cases in Bangkok [5] showed spatial dependence in both homotypic (same serotype) and heterotypic (different serotype) dengue cases at different time scales, reflecting complex interactions between local population immune profiles and dengue transmission. Local mosquito population dynamics, and human population density, immunity and mobility [7, 8, 9] are understood to be key determinants of these patterns. Peridomestic space spraying of insecticide is a mainstay of dengue control efforts in endemic settings, and the observed focal clustering of dengue cases provides a rationale for the common approach of targeted reactive insecticide spraying around the immediate neighbourhood of one or more notified dengue cases. The temporal scale of focal dengue transmission is important here, however, with some studies finding serological evidence for clustering of recent DENV infections but no excess of acute, prospectively-detected DENV infections [10], suggesting limited opportunity for reactive efforts to interrupt chains of transmission after detection of an index case. A lack of evidence for the efficacy and optimal implementation of conventional approaches to Aedes control [11, 12] together with the challenge of sustaining these activities at scale and over the long term [13], helps explain the ongoing occurrence of dengue outbreaks worldwide in spite of the efforts of vector control programs.

An alternative approach to the control of Aedes-borne diseases uses *Wolbachia*, a naturally occurring bacterium that is common in insect species but absent from *Ae. aegypti. Wolbachia* (*w*Mel-strain) infection of *Ae. aegypti* has been shown in the laboratory to reduce their transmission potential for dengue, chikungunya, Zika and Yellow fever viruses [14, 15, 16, 17, 18], and accumulating field evidence from randomized and non-randomized *w*Mel deployments demonstrates a significant reduction in the incidence of dengue and other Aedes-borne diseases in communities where *w*Mel has been established at a high level [19, 20, 21, 22, 23]. In a recent randomized controlled trial in Yogyakarta, Indonesia, (the ‘Applying *Wolbachia* to Eliminate Dengue’ [AWED] trial), the incidence of virologically-confirmed dengue cases was 77% lower in neighbourhoods where *w*Mel was successfully introgressed into local *Ae. aegypti* compared to areas that did not receive *w*Mel deployments [23]. The cluster randomized design of the AWED trial, with *w*Mel deployment into 12 of 24 contiguous clusters (average area 1 km^2^) in a highly urban study setting, means that movement of both humans and mosquitoes between treated and untreated clusters may have biased the measurement of the *w*Mel intervention effect toward the null [24, 25]. In a per-protocol analysis of the AWED trial accounting for participants’ time spent in the cluster of residence and other visited clusters, and the measured *w*Mel prevalence in those clusters, protective efficacy increased with incremental increases in estimated *w*Mel exposure, but never exceeded the intention-to-treat efficacy based on the treatment assignment of the cluster of residence [23]. This supports the home as a primary site of DENV exposure but the per-protocol analysis, as designed *a priori*, could not provide further insights into the transmission dynamics of the detected dengue cases, and specifically the extent to which dengue cases detected in residents of *w*Mel-treated clusters represented local transmission versus DENV infections acquired in untreated areas of Yogyakarta city.

Here we use spatiotemporal clustering analyses to explore the transmission-relatedness of dengue cases detected in *w*Mel-treated and untreated areas of Yogyakarta city, and the extent to which human movement between treated and untreated clusters may have biased the measurement of the *w*Mel intervention effect [25]. We use the geolocated residences of virolgically-confirmed dengue cases and test-negative controls enrolled in the AWED trial to test the hypotheses that dengue cases in Yogyakarta cluster in space and time in the absence of *w*Mel, and that this clustering is disrupted in *w*Mel-treated areas.

## 2 Results

### 2.1 Spatial and temporal distribution of dengue cases and test-negative controls

We examined the spatial and temporal distribution of 385 virologically-confirmed dengue cases and 5,921 participants with dengue test-negative febrile illness (test-negative controls) enrolled in the AWED cluster randomized trial of *w*Mel in Yogyakarta, Indonesia. Among the enrolled participants, those with test-negative illness were distributed throughout the entire 27-month study period in both intervention and untreated arms, with a peak in the first quarter of 2019 (Figure 1A). The majority of dengue cases in both study arms were enrolled during the wet season (January - May) in 2019 and 2020 (Figure 1B), with very few cases detected in the first year of trial enrolment in either study arm. As previously reported [23], amongst the 385 dengue cases, only 67 (17%) were resident in one of the 12 *w*Mel-treated clusters and the remaining 318 (83%) were resident in untreated clusters (Figure 2A). All four dengue virus (DENV) serotypes were detected, with a predominance of DENV2 (40%) and DENV4 (23%) (Supplementary Table 1 and Figure 2A). Fourteen dengue cases in the intervention clusters and 53 in the untreated clusters had indeterminable DENV serotypes and were therefore excluded from the spatiotemporal analysis, as the measure used to infer transmission-relatedness relies on the identification of homotypic dengue case pairs.

**Figure 1:**
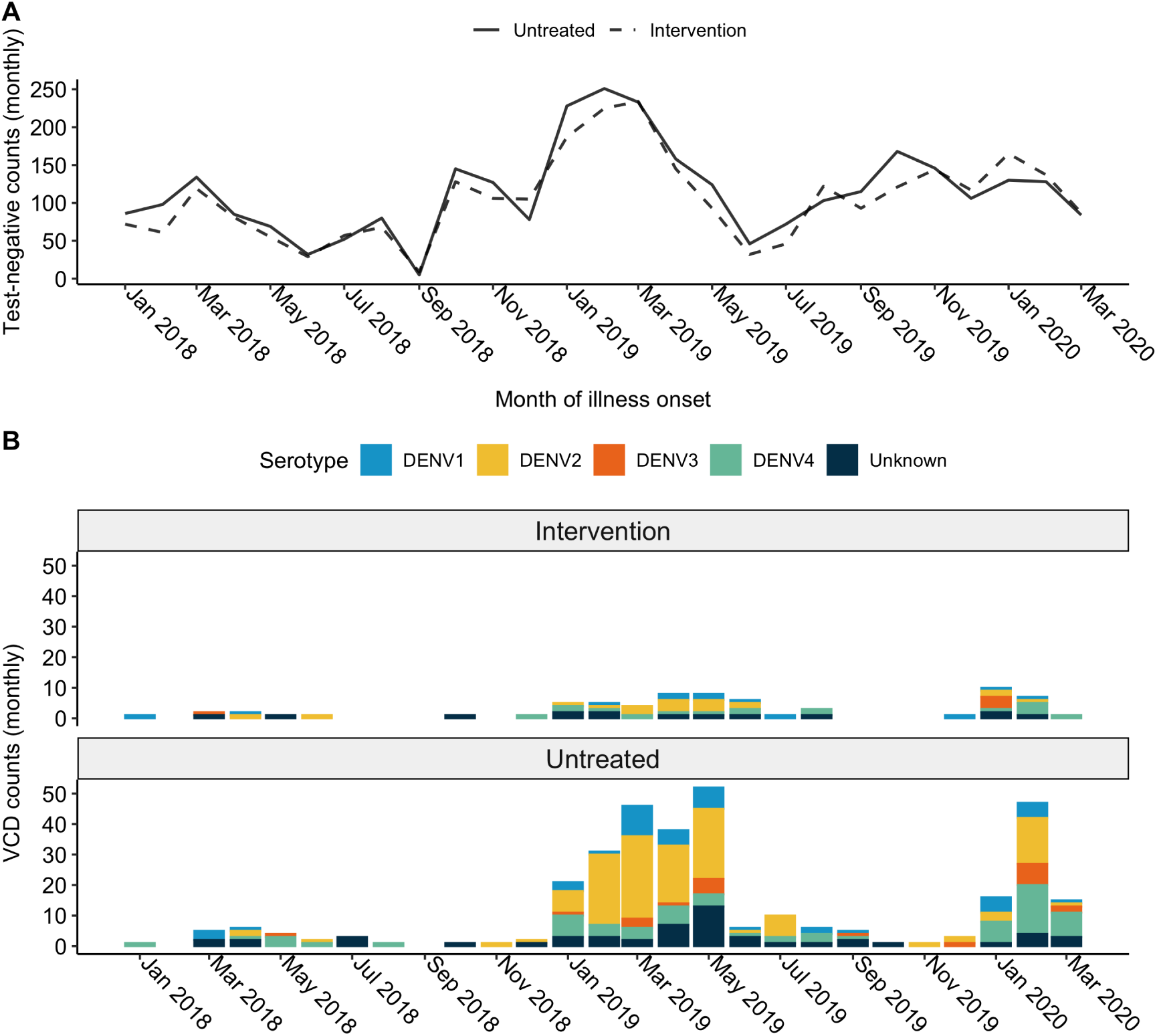
Time series plot for illness onset among A) test-negative controls and B) virologically-confirmed dengue cases included in the primary analysis of the AWED trial in Yogyakarta, Indonesia from January 2018 until March 2020, by intervention arm. No dengue cases were enrolled in September 2018 and, in accordance with the trial protocol [26], the test-negatives enrolled during that month were excluded from the analysis dataset.

**Figure 2:**
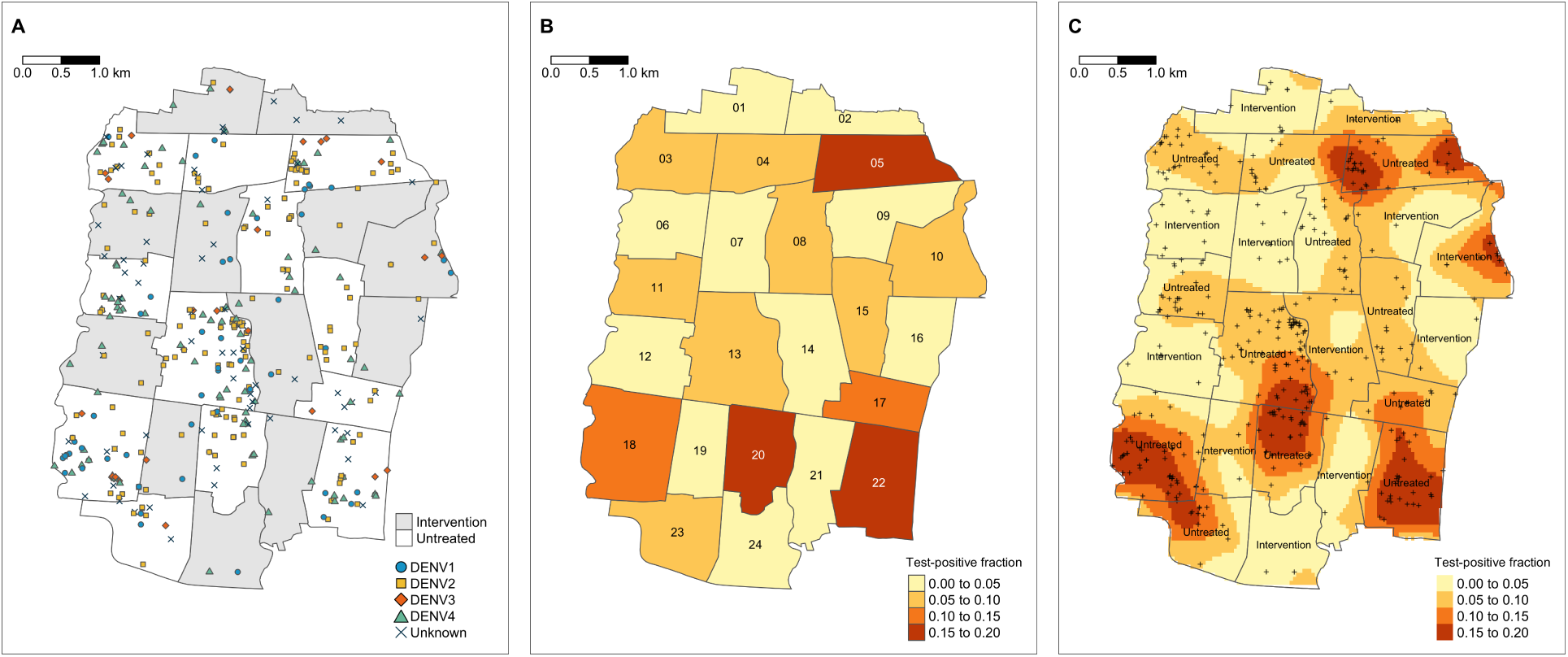
Spatial distribution of A) enrolled dengue cases by serotype across Yogyakarta City, B) the cluster-aggregate test-positive fraction, i.e., the proportion of enrolled dengue cases among the total number of individuals enrolled in each cluster, and C) kernel smoothing estimates of the spatially-varying test-positive fraction. Each map includes participants enrolled from January 2018 through March 2020. The borders in each map represent the cluster boundaries for the AWED trial. Clusters are labelled with their numerical code in panel B and their intervention status in panel C. Points represent the geolocated households of virologically confirmed dengue cases. Areas with darker shading in panels B and C have a higher proportion of dengue cases among enrolled AWED participants than areas with lighter shading. Smoothing bandwidth was selected by cross-validation.

Among the 5,921 participants with test-negative illness, 2,838 (48%) were resident in *w*Mel-treated clusters and 3,083 (52%) in untreated clusters. The map in Figure 2B incorporates the test-negative controls to account for the spatial distribution of the underlying healthcare-seeking population that gives rise to the enrolled dengue cases. Each cluster is shaded according to the cumulative test-positive fraction, i.e., the proportion of enrolled individuals who tested positive for virologically confirmed dengue in each cluster over the 27 months of enrolment. The cluster-specific test-positive fractions ranged from 5.9% to 23.8% in the 12 untreated clusters, whereas the test-positive fraction was ≤3.7% in 11 of the 12 intervention clusters. The one exception is intervention cluster 10, on the north east boundary of the study site, which had among the lowest overall enrolment rate of all clusters, with 87 test-negative controls and eight dengue cases enrolled (8.4% test-positive fraction). Six of eight dengue cases resided within 50m of the external boundary.

For a more granular view of the spatial distribution of dengue cases and test-negative controls, kernel smoothing was used to visualize the spatially-varying test-positive fraction (Figure 2C). Consistent with the cluster-level representation, the highest proportions of dengue case occurrence (aggregate over the 27-month trial period) fall within untreated clusters, and these areas of heightened dengue enrolment extend across the boundaries between untreated clusters.

### 2.2 Impact of *w*Mel on spatial dependence

Understanding the dynamics of DENV transmission in the absence and presence of *w*Mel is of primary interest, but directly inferring chains of transmission is difficult. We instead assume that pairs of homotypic (same serotype) dengue cases enrolled in the AWED study with illness onset within a pre-specified number of days of each other are potentially transmission-related, whereas pairs of heterotypic (different serotype) dengue cases or test-negative controls within the same space-time window are assumed not to be transmission-related. We compare the spatial distributions of these two populations of participant pairs as an indicator of spatiotemporal dependence in DENV transmission.

To assess small-scale spatiotemporal clustering in dengue cases throughout the trial area, we employ a global measure of spatiotemporal clustering, *τ* [27]. This measure captures the overall tendency of homotypic dengue cases to occur within specified space-time windows *above and beyond* that observed in the enrolled study population due to secular factors such as healthcare-seeking behaviour and environmental conditions (*Methods*). The numerator of this ratio-based estimator identifies, among those enrolled within a particular space-time window, the number of serotype-homotypic dengue pairs relative to the number of pairs of enrolled individuals who are assumed not to be transmission-related; the latter group includes the test-negative controls as well as any heterotypic dengue pairs. As such, the numerator of the *τ* -statistic is akin to an estimate of the odds of observing a homotypic dengue pair among all enrolled pairs in a given space-time window. The denominator of the *τ* -statistic is constructed the same way, but without restriction on the spatial window. Therefore, *τ >* 1 indicates that two enrolees are more likely to be homotypic dengue cases if they fall within the specified space-time window than if they fall anywhere across the study area. While we will refer to the *τ* -statistic as an odds ratio, there are subtle differences in this estimator that do not make it equivalent to the standard epidemiological odds ratio parameter.

The estimator is first applied to the overall study area, naive to local *w*Mel intervention status, with a time window of 30 days. Given that the intervention and untreated clusters are interspersed across the city, this serves as a global test to identify whether there is any evidence of spatial dependency in dengue cases overall before disaggregating by intervention arm. We find evidence of spatial dependence among homotypic cases occurring within 30 days at distances up to 300m, with the greatest relative odds of homotypic cases occurring within 50m of an index case (Figure 3). The odds that an individual enrolled within 30 days and 50m of an index case is a homotypic dengue case is 10.5-fold (95% CI: 5.6, 17.5) higher than for any individual with illness onset occurring within 30 days across the entire study area.

**Figure 3:**
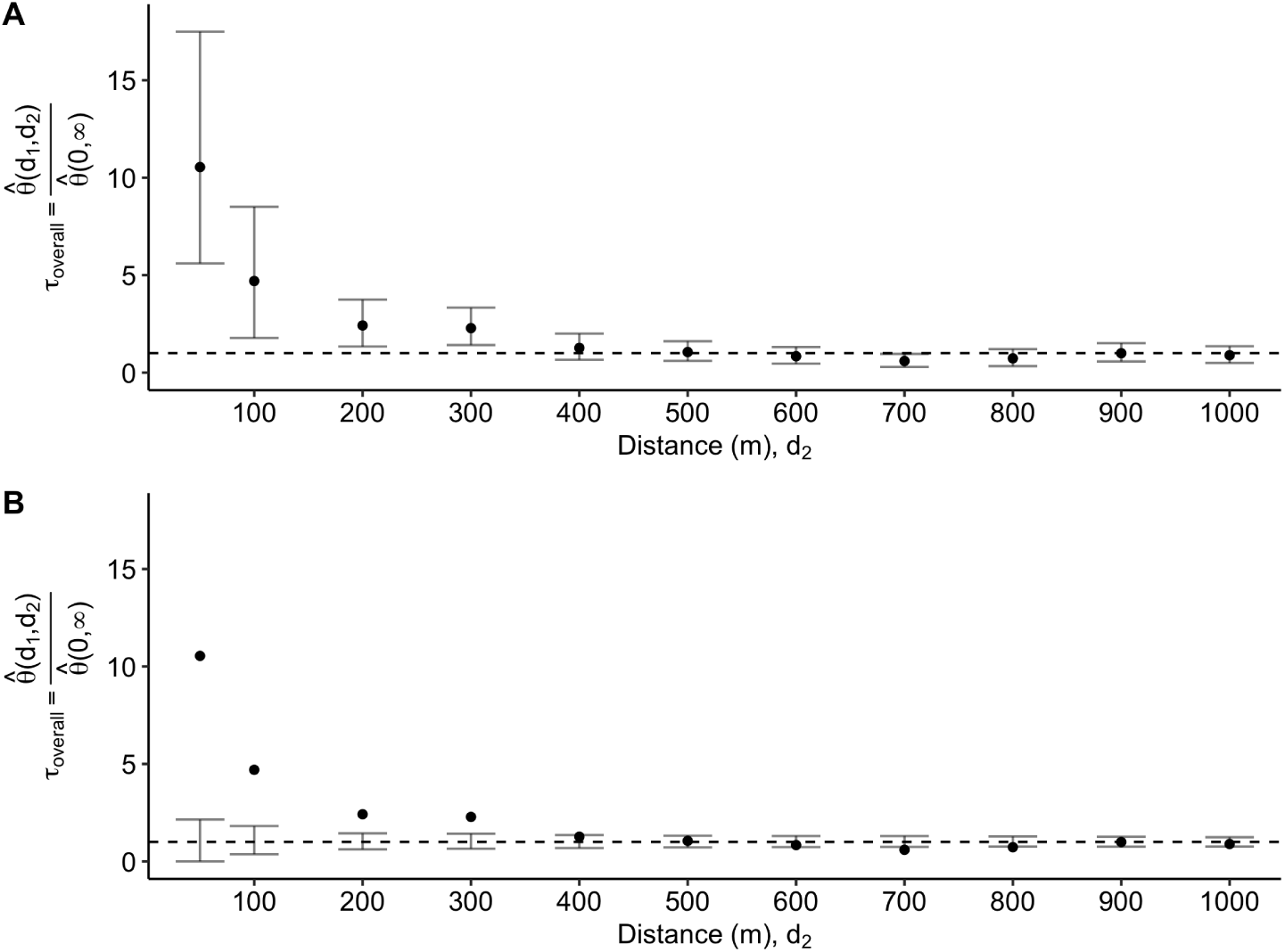
Estimated odds ratio, *τ*, comparing the odds of a homotypic dengue case pair within (*d*_1_, *d*_2_) versus the odds of a homotypic dengue case pair at any distance across the entire study area among participant pairs with illness onset occurring within 30 days. Variability in estimation is visualized in two distinct ways. A) displays the pointwise 95% confidence interval (CI) based on 1,000 bootstrap resamples of the data and B) shows the pointwise 95% CI on the permutation-based null rejection region based on 1,000 permutations of the data.

We then employ the *τ* -statistic to compare whether this spatiotemporal clustering in dengue cases differs between the untreated and *w*Mel-treated intervention arms by considering each of the 24 randomized clusters as independent units and estimating the small-scale spatiotemporal dependence of homotypic dengue cases within each cluster. Figure 4 shows the cluster-level estimates of the *τ* -statistic, together with the arm-level estimates of spatial dependence (calculated via a modified geometric mean: *Methods*). Statistical significance is inferred by comparison of the point estimate with the 95% CI on the permutation-based null distribution (bars) from 1,000 permutations of the data. The cluster-specific results highlight the sparsity of homotypic case pairs in the intervention arm. Six of the twelve clusters randomized to the intervention arm (clusters 2, 9, 16, 19, 21, 24) and only one of the twelve untreated clusters (cluster 17) had no dengue case pairs enrolled within 30 days of each other and with the same infecting serotype (ie. potentially transmission-related). As such, the *τ* -statistic is inestimable within these clusters.

**Figure 4:**
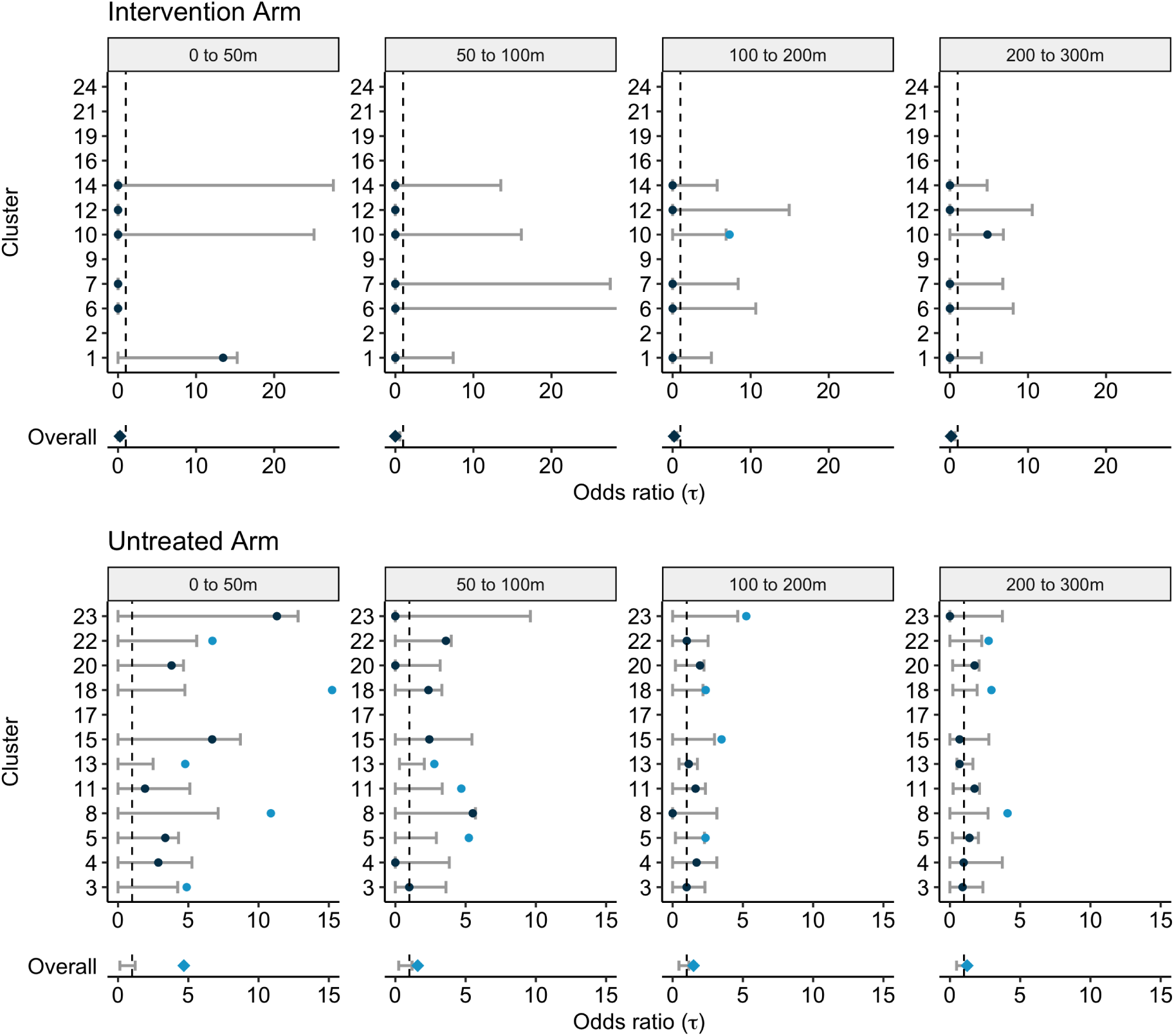
Cluster-specific and pooled arm-level estimates of the *τ* -statistic (points) and 95% CIs on the null distribution (error bar) generated from 1,000 simulations where the geolocation of participants is randomly permuted within each cluster. Each panel displays the estimated spatial dependence for homotypic case pairs with illness onset occurring within 30 days and resident within a given distance interval (meters) from each other. Estimated spatial dependence that is inconsistent with the null hypothesis is present when the point estimate falls outside of the 95% CIs of the null distribution and, for improved visibility, is marked by the light blue points. The overall point estimate for each trial arm is found by taking the geometric mean of the cluster-level estimates and is then compared against the 95% CIs of the null distribution of the permuted geometric mean.

Even when there is data to support estimation, the estimated pointwise 95% CIs of the permutation-based null distributions produce expansive limits. In all but one of the clusters where wMel-infected mosquitoes were deployed, the observed spatiotemporal dependence is consistent with the null hypothesis of no spatiotemporal dependence. Visually, this corresponds to the point-estimate from the observed data falling within the confidence bands on the permutation-based null distribution. The one inconsistent result in the intervention arm is the estimated spatiotemporal dependence in cluster 10 based on homotypic case pairs occurring within 30 days and 100m to 200m. It is worth noting that there were only two pairs of homotypic dengue cases (one pair with DENV1 and one pair with DENV3) observed within 30 days of each other in cluster 10. In contrast, five of 11 untreated clusters returned point estimates of spatiotemporal dependence within 50m of an index residence that appear inconsistent with the null hypothesis of no spatiotemporal dependence. For the untreated arm overall, spatiotemporal dependence was estimated to exceed what would be expected under the null hypothesis up to a distance of 300m, with the greatest relative odds of a homotypic case occurring for enrolled individuals with residences within 50m of an index case (*τ* (0, 50) = 4.7; 95% CI of the null distribution: 0.1, 1.2).

The disruption of spatiotemporal clustering of dengue cases by *w*Mel is underscored in Figure 5, which shows the geolocated residences of the 160 dengue cases that had illness onset within 30 days and residence within 300m of another case with the same infecting serotype (ie. homotypic case pairs). In Figure 5, unlike in the cluster-level *τ* -statistic analysis, cluster boundaries have been ignored so that all homotypic case pairs within the given space-time window are displayed. The vast majority of these potentially transmission-related dengue cases fall within the untreated regions. As described previously, only two case pairs fall fully in an untreated cluster, namely cluster 10.

**Figure 5:**
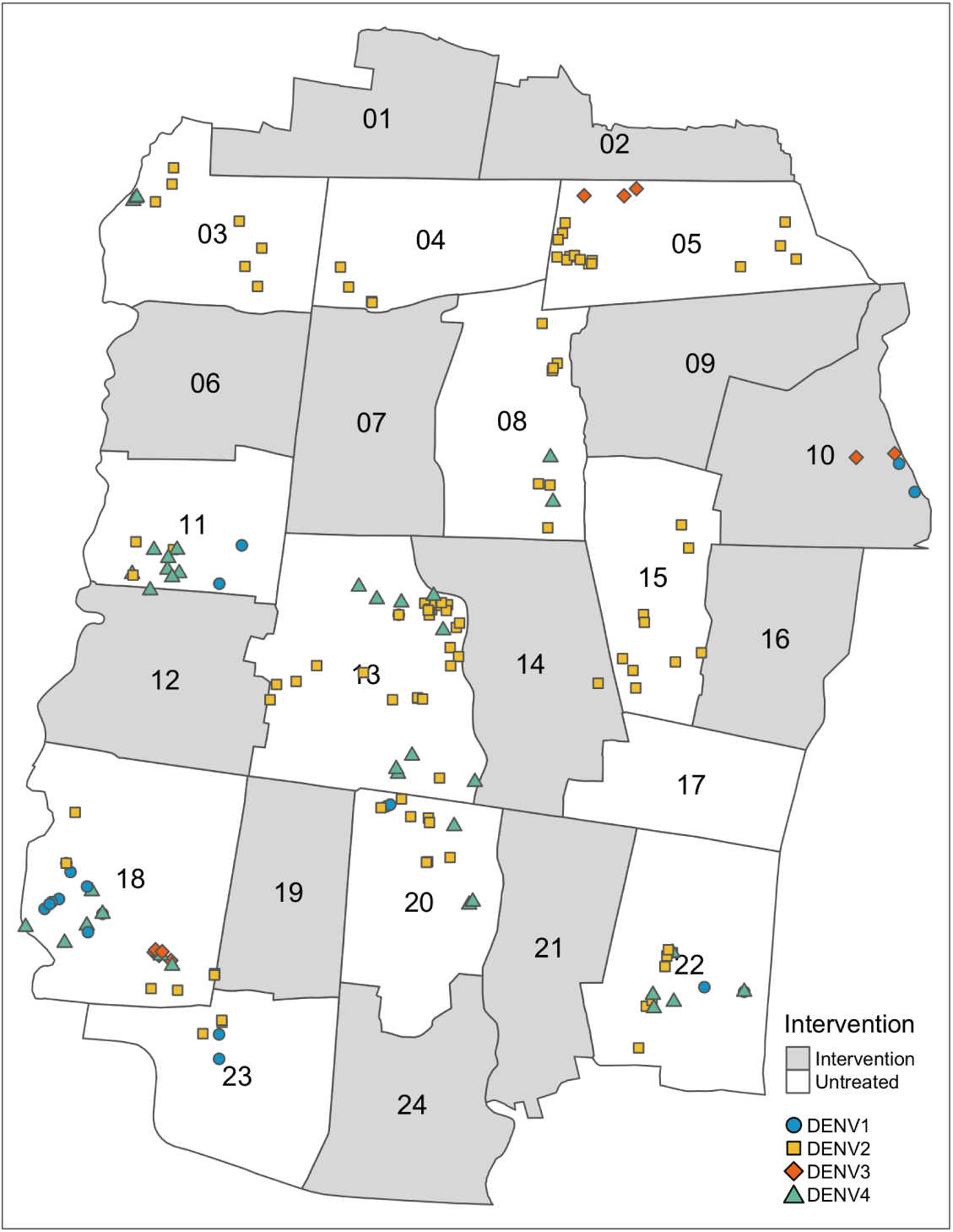
Residential locations of the 160 enrolled serotyped dengue cases involved in homotypic pairs with residences within 300m and illness onset within 30 days, including pairs that cross cluster boundaries.

### 2.3 Sensitivity analysis

The test-negative controls were enrolled after presenting with acute febrile illness and, as such, may exhibit their own spatial structure beyond that of the underlying healthcare-seeking population. As a sensitivity analysis, we performed the overall *τ* -statistic estimation excluding the test-negative controls, instead relying on the ratio of homotypic to heterotypic dengue case pairs occurring within space-time windows to estimate spatial dependence. There was no meaningful difference observed when test-negatives were included or excluded from estimation (Figure S1).

We additionally performed sensitivity analyses to account for potential contamination effects for residences near cluster boundaries and to examine the effects of applying differing temporal clustering windows. The results are presented in Figure 6 where each panel displays the modified geometric mean and permutation-based 95% CIs of the null distribution of the geometric mean. The “Full data” panels display the same information as the “Overall” rows of Figure 4 and are included for ease of comparison with the sensitivity results. When individuals residing within 50m of any cluster boundary are excluded from the analysis, the magnitude of the estimated *τ* -statistic in the untreated arm decreases, but remains larger than expected under the null hypothesis of no spatial dependence up to a distance of 300m, as observed in the primary analysis. When the window of time between illness onset of case pairs was decreased from 30 days to seven days, there is evidence of spatial clustering of homotypic dengue cases up to 50m, and potentially from 100-200m, in the untreated arm. Finally, when an temporal interval of 2 weeks between illness onset of case pairs is used (the most likely serial interval between successive dengue illnesses[4]), there is evidence of spatial dependence up to 200m in the untreated arm. There remains no evidence of small-scale spatiotemporal clustering of dengue cases in the intervention arm under any of these sensitivity settings.

**Figure 6:**
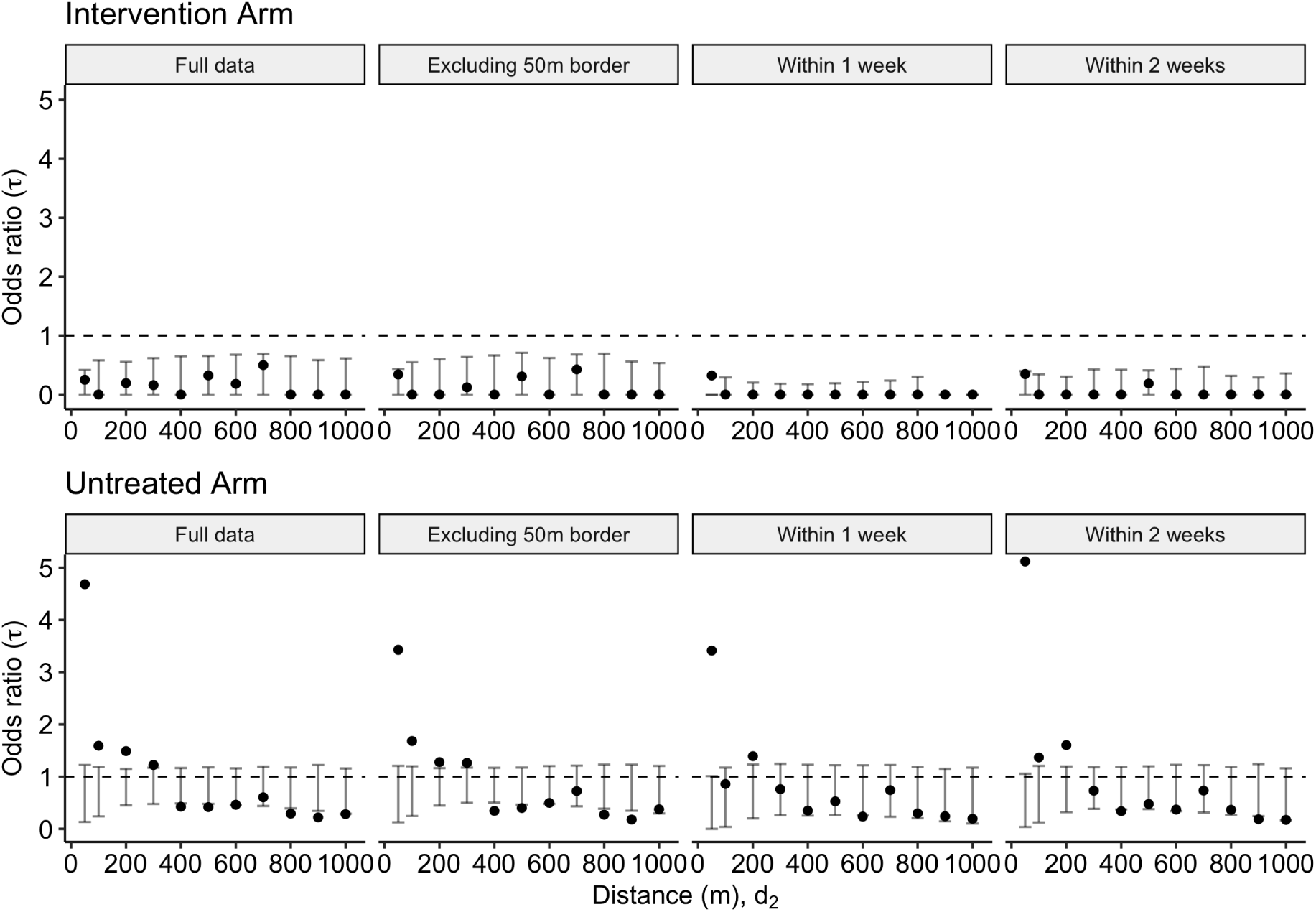
Sensitivity analyses and comparison with the primary analysis. Modified geometric mean odds ratio, *τ*, comparing the odds of a homotypic dengue case pair within the distance interval (*d*_1_, *d*_2_) versus the odds of a homotypic dengue case pair at any distance across the entire study area for 1) the full dataset, 2) the dataset excluding those within 50m of a cluster border, 3) participant pairs with illness onset occurring within 1 week of each other, and 4) participant pairs with illness onset occurring within 2 weeks of each other. The error bars denote the 95% CI on the null distribution generated by 1,000 permutations of the data.

## 3 Discussion

We show here that successful introgression of *w*Mel into the local *Ae. aegypti* mosquito population disrupts the focal transmission of dengue virus. The intention-to-treat analysis of a cluster randomised controlled trial of *w*Mel deployments in Yogyakarta, Indonesia previously reported a 77% reduction in dengue incidence in *w*Mel-treated areas [23]. However, the inability to account fully in the primary analysis for human and mosquito movement between treated and untreated clusters may have biased the efficacy estimate towards the null, making the reported protective efficacy of *Wolbachia* an underestimate[25]. The findings reported here from a secondary analysis of the trial data demonstrate an absence of spatiotemporal clustering among the 53 serotyped dengue cases detected in *w*Mel-treated areas. These results are consistent with the hypothesis of an even larger *w*Mel intervention effect than measured in the primary analysis of the AWED trial, and raise expectations that area-wide coverage of Yogyakarta with *w*Mel could result in near elimination of local DENV transmission.

In six of the twelve clusters where *w*Mel deployments occurred, no homotypic dengue case pairs occurred in any 30-day window throughout the 27 months following *w*Mel releases. Among the remaining six *w*Mel-treated clusters where the spatial dependence of homotypic dengue case pairs was estimable, only one cluster on the northeast border of the trial site had any homotypic dengue case pairs that could plausibly have been transmission-related. In contrast, in untreated areas we found evidence of clustering in dengue incidence within space-time windows of 30 days and 300m, with increasing spatiotemporal dependence within decreasing intervals of space (up to 50m) and time (up to 7 days). The lack of spatiotemporal clustering among the dengue cases resident in *w*Mel-treated areas of Yogyakarta does not preclude the possibility that infection occurred in the cluster of residence, and that other transmission-related infections went undetected or were asymptomatic. However, since the sensitivity of dengue case detection can be expected to be equivalent, on average, between treated and untreated arms of the AWED trial, these findings strongly suggest that the dengue cases in the intervention clusters may have acquired their infection outside of their cluster of residence. Information collected in the AWED trial on the travel history of participants during the ten days prior to illness onset, together with planned genomic analysis of DENV detected in trial participants, will support a more direct assessment of the potential transmission-relatedness between dengue cases detected in intervention clusters and those resident elsewhere in the trial area.

The distortion by *w*Mel of spatial patterns in dengue case occurrence is evident in the visualizations of the dengue case time series aggregate over 27 months, even before considering the two key components in potential transmission-relatedness: serotype and case onset date. At an aggregate area level, despite being completely intermixed with the untreated clusters, the cluster-specific test-positive fraction estimates for *w*Mel-treated clusters are all considerably lower than the test-positive fractions in the untreated clusters, with the exception of intervention cluster 10. The results of the kernel estimator, which does not differentiate between serotype or take artificial study boundaries into consideration, suggest that the areas where dengue test-positive febrile illness is highest are located in untreated clusters and tend to extend across borders with other untreated areas rather than borders with intervention clusters. When serotype and case onset date are accounted for via the spatial-temporal clustering *τ* -statistic, this observation is strengthened further.

The fine scale spatial clustering of dengue cases in untreated areas of Yogyakarta closely mirrors findings reported by others from Vietnam [6], Thailand [4, 6, 5], Australia [3], Peru [9], and Taiwan [28] despite differences in analysis methods, ecological setting and human population characteristics. We observed strongest clustering of dengue cases within 50m and 7 days, declining with increasing spatial and temporal distance, which is supportive of focal DENV transmission within households and the immediate neighbourhood in the absence of *Wolbachia*. Similar focal clustering has been reported from Thailand, where the authors found strongest evidence of clustering at a 15-17 day interval and distances less than 200m [4]. These spatiotemporal patterns provide the rationale for the common approach of applying conventional vector control interventions, primarily insecticide spraying, in a reactive manner around the households of notified cases that cluster in space and time. In Yogyakarta, notification of hospitalised dengue cases to the District Health Office with evidence of local transmission (i.e. additional confirmed or suspected dengue cases within 2 weeks and 100m) will usually trigger perifocal insecticide fogging, with malathion most commonly used in recent years, and application of a larvicide such as pyriproxyfen to water containers. Insecticide resistance and insecticide penetration to indoor resting places is a challenge, and the limited ability of these approaches to meaningfully impact dengue virus transmission is evidenced by the high baseline dengue burden and paediatric seroprevalence in Yogyakarta [29], and recurring dengue outbreaks in other locations where large efforts are expended on Aedes vector control [30].

One of the strengths of the *w*Mel method for dengue control is its efficacy against the four DENV serotypes [23]. However, this results in very little case data for the estimation of spatiotemporal clustering. Only 67 virologically confirmed dengue cases were enrolled in the intervention clusters throughout the 27-month AWED study, of which, only 53 had identifiable DENV serotypes. When examining the spatiotemporal dynamics of transmission, the lack of potentially transmission-related dengue is itself a critical finding, despite being difficult to represent statistically. The completion of *w*Mel deployments throughout the AWED untreated clusters in January 2021 is expected to result in an even greater impact on dengue in Yogyakarta in coming years, and even raises the prospect of local elimination. Monitoring progress towards dengue elimination in Yogyakarta will require the development of an appropriate surveillance framework and statistical methods for demonstrating absence of disease, which differ from those used when disease is present [31].

A considerable hurdle in spatiotemporal analyses is the identification and geolocation of the underlying population at risk. Cross-sectional surveys are of limited utility for examining spatiotemporal patterns in the occurrence of self-limiting acute infections like dengue, as virological markers of acute infection are short-lived and detection of antibody is not informative about the timing of infection. Prospective cohort studies with clinical or serological endpoints are expensive, time-consuming and logistically complicated to carry out, and their sensitivity for detection of case clustering may be limited unless very large. As such, many studies rely on passively collected case data and compare these counts against “total population” census estimates, introducing room for bias when the total population is not the true population at risk because it includes individuals with immunity as well as those whose healthcare-seeking behaviour may preclude their detection in facility-based data sources. The test-negative design provides a new framework for simultaneously sampling cases and controls from the underlying at-risk, healthcare-seeking population. Methodological research is in progress to further explore the benefits of this design for spatiotemporal analyses of infectious disease.

Further research and development of the small-scale spatial dependence estimator *τ* is underway and will be helpful for future applications [32]. In this work, we have attempted to follow the recommended best practices when applying *τ* in a diagnostic manner [33]. In the context of the test-negative design specifically, *τ* also requires further methodological assessment. The authors of the method have pointed out that both the numerator and the denominator are dependent on the spatiotemporal distribution of cases, and importantly, controls [27, 5]. The controls in the AWED study may not be a random sample from the underlying spatial distribution of the population. If there are other processes driving spatial dependence in the test-negatives other than the spatial distribution of the underlying population (e.g., transmission of other pathogens), then this would affect the calculation and interpretation of the *τ* -statistic. As such, the test-negatives more likely represent a random sample from the care-seeking population. The estimates of the *τ* -statistic should then be interpreted as the spatial dependence in homotypic dengue cases *over and above* any spatial patterns in the underlying health-care seeking population that gave rise to the dengue cases. However, as demonstrated in Figure S1, the conclusions of spatial dependence do not change upon the exclusion of test-negatives from the estimation process.

Dengue was a relatively rare outcome in the AWED study sample, present in only 6% of individuals in the analysis set. As such, using bootstrap resampling as the basis for statistical inference resulted in two complications. First, previous work has demonstrated that blocked bootstrap resampling of spatial data is generally a more appropriate approach to inference of spatial estimators as it retains a level of spatial correlation among observed case locations that is lost when resampling individuals [34]. The performance of the blocked bootstrap has yet to be explored in the context of the *τ* -statistic for small-scale spatial dependence. Second, resampling with replacement at the individual-level when there are overwhelmingly more negative controls than cases results in some bootstrap resamples devoid of any cases. Given the manner in which cases and controls are ascertained in the test-negative design, resampling conditional on outcome is a problematic solution to this problem. Instead, we rely on permutation methods for assessing whether the observed spatiotemporal dependence is consistent with what would be expected under the hypothesis of no spatiotemporal dependence, which only requires a reshuffling rather than a resampling of the dataset [27]. Further methodological development in estimation and inference for the *τ* -statistic is needed in the rare-disease, test-negative design setting.

The current work examines DENV transmission under a binary intervention status based on household residence. This serves as only a proxy of an individual’s true intervention experience. The presence of *w*Mel in the monitored mosquito populations was strikingly homogeneous within the intervention areas, but, by the second year of the trial, was also detected at the edges of the untreated cluster borders [23]. Additionally, human movement could affect an individual’s risk of infection and transmission [9]. AWED participants’ geolocated movements over the ten days prior to illness onset were recorded at enrolment, providing a unique opportunity to gain further insight into the extent of human mobility and its role in transmission. Incorporating such fine-scale spatial and temporal data will allow for the investigation of DENV transmission beyond the proxies of geolocated residence and intervention assignment.

This work provides the first report of spatiotemporal clustering of dengue in Yogyakarta in the absence of *w*Mel, replicating others’ reports from multiple endemic settings. Importantly, it shows that in areas randomly allocated to *w*Mel deployments, the sustained introgression of *w*Mel into the local *Ae. aegypti* population successfully disrupts the focal transmission of dengue virus, consistent with a protective efficacy of *Wolbachia* that may be greater than previously reported.

## 4 Methods

### 4.1 Data source

Data was collected during the Applying *Wolbachia* to Eliminate Dengue (AWED) trial (NCT03055585) – a parallel, two-arm, cluster-randomized test-negative design study carried out in Yogyakarta, Indonesia from January 2018 to March 2020. The trial design and results have been described elsewhere [26, 23]. Written informed consent for participation in the clinical component of the trial was obtained from all the participants or from a guardian if the participant was a minor. In addition, participants 13 to 17 years of age gave written informed assent. The trial was conducted in accordance with the International Council for Harmonisation guidelines for Good Clinical Practice and was approved by the human research ethics committees at Universitas Gadjah Mada and Monash University [26]. Briefly, the city was divided into 24 contiguous clusters, twelve of which were randomly assigned to receive *w*Mel-infected *Ae. aegypti* releases. Routine vector control activities continued throughout the study area. Individuals 3 - 45 years old presenting to government primary care clinics with acute febrile illness who were resident in the study area, had no localising symptoms suggestive of a non-dengue diagnosis, and had not been enrolled within the previous 4 weeks were invited to enrol and their residence and places visited during 10 days prior to illness onset were geolocated. A positive result in either dengue PCR or NS1 antigen ELISA distinguished virologically-confirmed dengue cases from test-negative controls (negative in DENV PCR, NS1 ELISA and IgM/IgG ELISA) and a subset of participants excluded from analysis (negative in PCR and NS1 but positive in IgM/IgG, or with inconclusive diagnostic results). The infecting serotype was determined for PCR-positive dengue cases.

### 4.2 Statistical Methods

#### 4.2.1 Global analysis of spatial dependence

To characterize the small-scale spatiotemporal dependence of homotypic dengue cases, we employ a global measure, the *τ* -statistic [35]. This measure captures the overall tendency of homotypic dengue cases to occur within specified space-time windows above and beyond that observed in the enrolled study population due to secular factors (e.g., healthcare-seeking behaviour and environmental conditions). This method compares the odds that an enrolled pair of individuals within a given time and distance, *θ*(*d*_1_, *d*_2_), are homotypic dengue cases against the odds that an enrolled pair of individuals within a given time but at any distance, *θ*(0, ∞), are homotypic dengue cases (Equations 1, 2). As such, a pair of enrolled individuals *i* and *j* are considered to be potentially transmission-related (*z*_*ij*_ = 1) if they have the same infecting DENV serotype and have illness onset within a specified time window, and are otherwise assumed to be non-transmission related (*z*_*ij*_ = 0). Specifically,

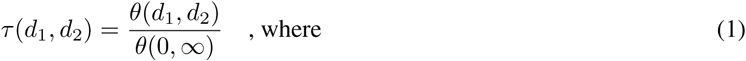

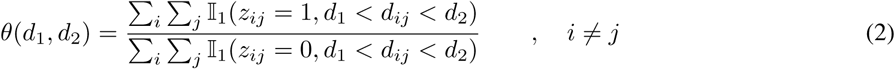

For Equation 2, the sums range over all test-positive and test-negative trial participants for the overall estimate of the *τ* -statistic. For the cluster-specific estimates, the sums range over all cluster-specific test-positive and test-negative trial participants.

Based on the observed spatial dependence in dengue cases reported previously in other settings, we examine the *τ* -statistic at 0-50 meters and 50-100 meters, and then 100 meter intervals up to a maximum distance of 1,000m from an index case.

Permutation-based null distributions from 1,000 reshuffles of the location data provided the basis of hypothesis testing used to evaluate statistical significance of the estimated *τ* -statistic. For the overall estimates of *τ* ignoring intervention assignment, this includes all enrolled test-negative controls and test-positive cases with determinable DENV serotype. For the cluster-specific estimates of *τ*, the location data of the cluster-specific test-negatives and test-positives are permuted within each cluster. Bootstrapped confidence intervals, obtained by 1,000 random resamples of the individual-level data, were obtained for the intervention-naive *τ* analysis. Bootstrapped confidence intervals for the cluster-specific estimates of *τ* are not provided, given the sparsity of specific serotype cases at the cluster-level. Arm-level estimates of the *τ* -statistic are based on a modified geometric mean procedure common in settings with high frequencies of zeroes; the value of one is added to each cluster-level estimate of *τ* before the estimation of the geometric mean and then subtracted from the exponentiated result [36].

### 4.3 Data Availability

The data underlying Figures 1, 2B, 3, 4 and 6 are accessible at the corresponding author’s GitHub repository (https://github.com/sdufault15/awed-spatial-temporal). Participants’ residential address was collected and stored under the ethical approval of the AWED trial protocol. Because the geolocated residence is considered personally identifiable information, individuals seeking access to this data should contact Katherine Anders to discuss obtaining ethical approval or accessing a deidentified individual-level version of the dataset.

### 4.4 Code Availability

All analyses were performed using R version 4.0.4 “Lost Library Book”. General data cleaning and management relied heavily on the “tidyverse” [37], “here” [38], “rgdal” [39], and “geodist” [40] packages. Map visualizations were constructed using “tmap” [41]. Kernel smoothing estimation was performed via “spatstat::relrisk” [42]. The code used for this analysis is available at the GitHub repository (https://github.com/sdufault15/awed-spatial-temporal) maintained by the first author.

## Data Availability

Participants' residential address was collected and stored under the ethical approval of the AWED trial protocol. Because the geolocated residence is considered personally identifiable information, individuals seeking access to this data should contact Katherine Anders to discuss obtaining ethical approval or accessing a deidentified version of the dataset.  All analyses were performed with R and the analytical code can be found on GitHub (https://github.com/sdufault15/awed-spatial-temporal).

## Acknowledgements

We thank Henrik Salje for his informative discussions around the use and interpretation of the *τ* estimator. We acknowledge the contribution of all investigators in the AWED Study Group to the implementation of the trial. We thank the leadership and residents of Yogyakarta for their support and participation in the trial. We gratefully acknowledge the Tahija Foundation as funders of the AWED trial, and the Wellcome Trust and the Bill and Melinda Gates Foundation, which provided financial support to the World Mosquito Program.

