## Supplemental Table and Figure for "Disruption of spatiotemporal clustering in dengue cases by *w*Mel *Wolbachia* in Yogyakarta, Indonesia"

### Supplementary Material

**Table S1.** The distribution of virologically confirmed dengue cases and test-negative controls by AWED study arm.

| | Intervention | Untreated |
|---|---|---|
| Test-negative controls | 2838 | 3083 |
| Dengue cases | 67 | 318 |
| DENV1 | 12 | 45 |
| DENV2 | 20 | 134 |
| DENV3 | 5 | 22 |
| DENV4 | 17 | 70 |
| Unknown | 14 | 53 |

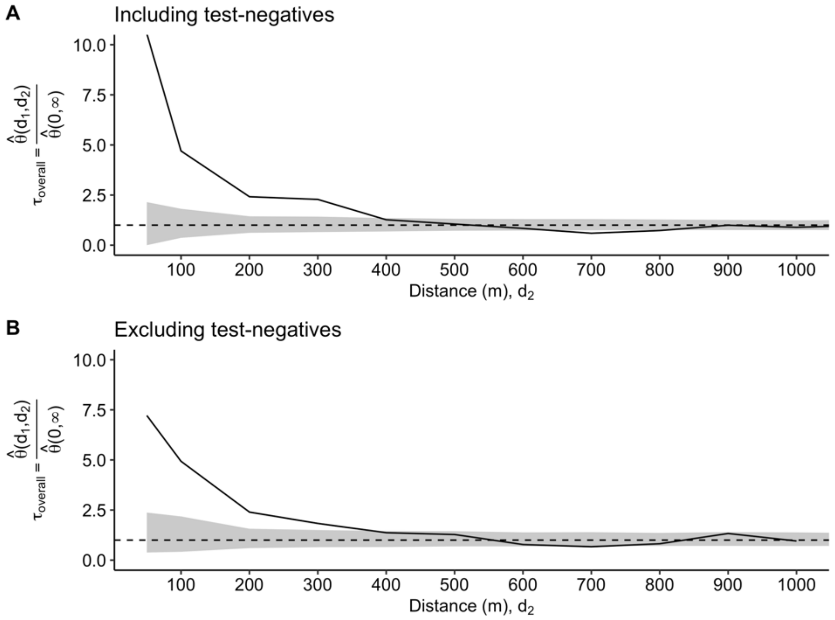


**Figure S1.** Estimated odds ratio, $\tau$, comparing the odds of a homotypic dengue case pair within ($d_1,d_2$) versus the odds of a homotypic dengue case pair at any distance across the entire study area with A) test-negative controls contributing to the counts of heterotypic pairs, and B) test-negatives excluded and based solely on the heterotypic and homotypic counts of dengue pairs. Variability in estimation is visualized in two distinct ways. A) displays the pointwise 95% confidence interval (CI) based on 1,000 bootstrap resamples of the data and B) sows the pointwise 95% CI on the permutation-based null rejection region based on 1,000 permutations of the data.
